# Effect of modified basic package of oral care on adolescents’ dental caries status in Copperbelt Province, Zambia; A Cluster Randomized Trial

**DOI:** 10.1101/2024.03.03.24303674

**Authors:** Severine N Anthony, Hawa S Mbawalla, Febronia K Kahabuka, Seter Siziya, Anne N Åstrøm

## Abstract

**Background:** Dental caries remains the major oral health challenge affecting more than half of adolescents globally. Most of the disease remain untreated, therefore, negatively impacting adolescents’ general health, well-being, and quality of life. Basic Package of Oral Care (BPOC) is a potential solution to the challenge, however, evidence on its effectiveness is scarce. This study primarily assessed the effects of applying modified BPOC on dental caries prevalence and secondarily on knowledge and behaviors related to dental caries among adolescents in Copperbelt Province, Zambia.

**Methods:** A parallel arms cluster randomized field trial (Reg-PACTR202210624926299) including 22 public secondary schools in Copperbelt province, Zambia, was carried out between January 2021 and March 2023. A pre-tested questionnaire was used to collect socio-demographics, knowledge and dental caries related behaviors data, while dental caries was assessed clinically using the caries assessment spectrum and treatment (CAST) at baseline and the follow-ups. The 1^st^ and 2^nd^ follow ups were conducted at 18-and 24months after baseline respectively. The analysis was based on intention-to-treat protocol using generalized estimating equations (GEE) and results are reported as OR (95% CI).

**Results:** Out of 1,794 participants at baseline, 1,690 (94.2%) and 1,597 (89.0%) were seen at 1^st^ and 2^nd^ follow ups respectively. A significant interaction (BPOC x time) for dental caries models at 18 months {OR (95%CI) = 1.3 (1.1, 1.6), p=0.003)} and 24 months {OR (95%CI) = 1.3 (1.1, 1.6), p=0.004)} was observed. Secondary outcomes with significant interactions included adequate knowledge models at 18 months {OR (95%CI) =1.5 (1.2,1.8), p<0.001} and 24 months {OR (95%CI) = 1.6 (1.3, 2.0), p<0.001} as well as use of fluoridated toothpaste twice or more per day at 18 months {OR (95%CI) = 1.6 (1.3, 2.1), p<0.001)} and 24 months {OR (95%CI) = 1.4 (1.2, 1.6), p<0.001)}. Subgroup analysis showed that the intervention group had better outcomes than the control group in terms of dental caries prevalence, adequate knowledge, use of fluoridated toothpaste twice or more per day, at 18- and 24 months.

**Conclusion:** The modified BPOC was effective in reducing prevalence of dental caries, improving knowledge on dental caries, and the frequency of using fluoridated toothpaste among Zambian adolescents. Further studies need to be conducted in order to address other factors affecting oral health related behaviors such as the school and home environment, social and cultural factors.

## Introduction

Despite dental caries being preventable, its prevalence continues to rise, especially in low- and middle-income countries [1]. A large proportion of dental caries remains untreated thus increasing demand for essential oral health services and on the other hand impacting negatively the individual general health and quality of life [2–4]. The World Health Organization (WHO) recommends to reduce the burden of oral diseases by emphasizing comprehensive system changes to shift from traditional curative approaches toward preventive approaches such as application of the basic package of oral care (BPOC) [5].

BPOC is geared towards provision of oral health interventions with proven effectiveness that are acceptable, practicable, and cheap to the majority of disadvantaged communities [6,7]. The strategy is critical to fulfilling the core goal of primary health care, which is to provide better health services to all. It is also in line with the World Health Organization’s (WHO) recommendation of reorienting traditional curative-oriented oral health services towards approaches that adhere to the principles of primary health care (PHC) [6,7].

BPOC consists of three packages: Oral Urgent Treatment (OUT), Atraumatic Restorative Treatment (ART), and Affordable Fluoride Toothpaste (AFT). OUT is achieved by offering simple extraction of teeth beyond restorable state and referral of complicated cases. ART is provided through restoring teeth with reversible pulpitis on cavities which can be accessed by hand instruments only without a need of rotary instruments then restored using glass ionomer cement. AFT can be achieved through regulatory strategies such as lowering or abolishing taxes on the items with the goal of reducing their market prices [6]. The World Health Organization recommends tailoring BPOC to the local environment and evaluating its effectiveness in improving oral health needs in various locations around the world [6].

Although the effectiveness of the individual components of BPOC has been demonstrated, evaluation of its overall success as a package has received little attention. A study in Cambodia demonstrated success in increasing extractions and ART restored teeth at follow up following BPOC intervention [8]. Another study reported success in reducing the incidence of early childhood caries among children of women who received BPOC during pre- and post-natal period [9].

No study reporting the effectiveness of BPOC as a package in preventing dental caries among adolescents was retrieved. This study is, therefore, aimed at assessing the effects of modified basic package of oral care on prevalence of dental caries among adolescents in Copperbelt Province, Zambia. The study secondarily assessed the effects of modified basic package of oral care intervention on knowledge, frequency of consuming sugary drinks and foods, frequency of use of fluoridated toothpaste per day and dental visits in the previous year. The hypothesis tested for the primary objective was; at follow ups the likelihood of dental caries occurrence is less in the intervention than in the control group. The hypotheses for the secondary objective were the likelihood of adequate knowledge, consuming sugary drinks and foods less than five times per day, using fluoridated toothpaste twice or more per day, and visiting a dentist at least once in the previous year are more in the intervention than in the control group.

## Methods

### Study design

The study used a parallel two arms cluster randomized controlled field trial design with one control group and one intervention group (allocation ratio 1:1). The trial was registered by Pan African Clinical Trial Registry (PACTR202210624926299) available at https://pactr.samrc.ac.za/TrialDisplay.aspx?TrialID=24046. CONSORT reporting guidelines for randomized trials (Supplementary file 1) have been adhered to in reporting all sections of this study [10,11]. The study commenced in January 2021 (recruitment of participants and collecting baseline data) and ended in March 2023 as illustrated in detail in the schedule of study activities (Supplementary file 2). The number of follow-ups was reduced as soon as the trial began from three initially intended, to two due to unanticipated school closures during COVID 19 pandemic.

### Ethical considerations

The study received ethical approval from three institutions: the study site (Tropical Diseases Research Centre, Zambia -IRB 00002911 FWA 00003729), training institutions (MUHAS Institutional Review Board, Tanzania -P. MUHAS-REC-4-2020-208), and the Regional Ethical Committee vest 191836, Norway). National Health Research Authority (permit no - NHRA00005/16/11/2020), provinvial health directorate and district education board secretaries granted permission to conduct the research. Written informed consent were obtained from parents or gurdians and participants were requested to sign a written assent form.

### Participants eligibility criteria and setting

All adolescents in grades eight to nine attending public secondary schools in Ndola, Masaiti and Mpongwe districts of the Copperbelt Province, Zambia were eligible to participate (Fig 1). The study excluded adolescents under active or passive orthodontic treatment.

**Figure 1:**
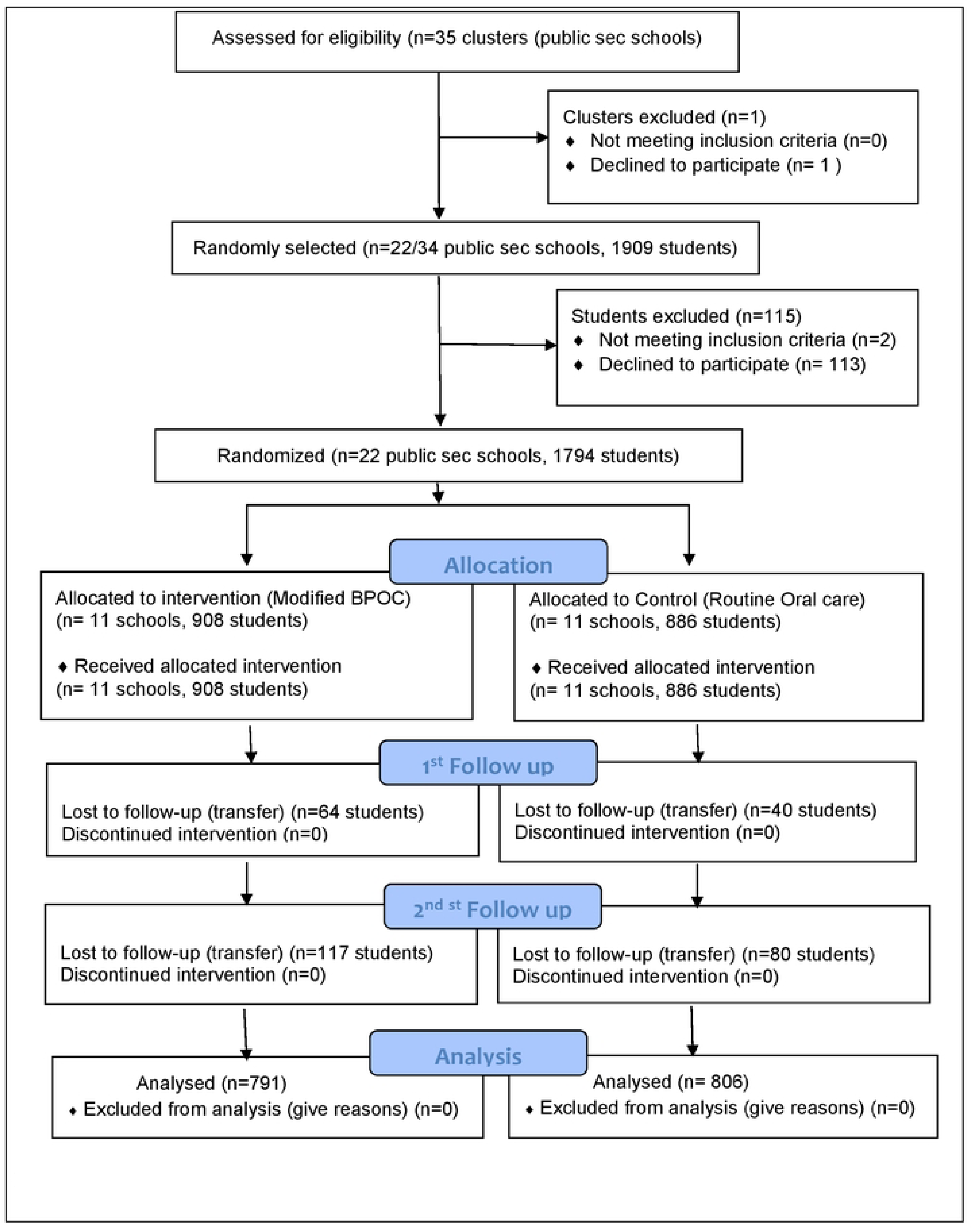
Flow diagram of the trial progress

**Figure 2:**
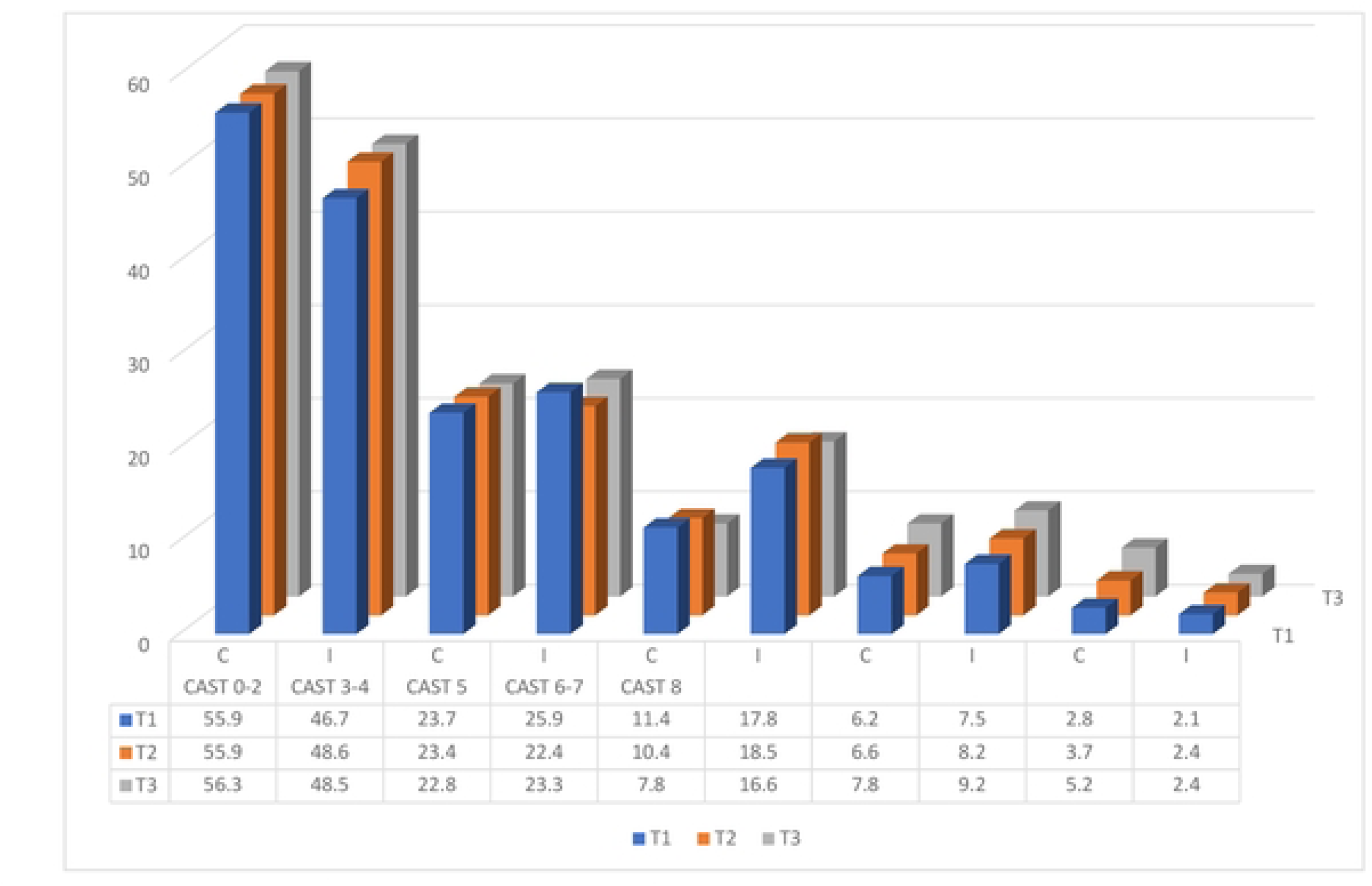
Trends in lhe distribution of dental caries clinical categories according to trial arms at baseline (T_1_), 6 months (1’_2_) and 12 months (T_3_)

## The trial intervention

### Experimental group

The participants in the experimental group received a modified World Health Organization - Basic Package of oral care (WHO-BPOC) for a period of six months in addition to their routine daily standard of oral care. The intervention adopted Oral Urgent Treatment (OUT) and Atraumatic Restorative Treatment (ART) components of the conventional WHO-BPOC and modified Affordable Fluoride Toothpastes (AFT) component. The conventional BPOC recommends effecting AFT component by lobbying for reduction of prices of toothpastes through government subsidies or tax reduction which could not be feasible in this study. In this study; AFT component was modified by providing each participant a 500g pack of fluoride toothpaste once every two months and a toothbrush once every three months in order to encourage future usage of the oral care products. Peer-Led Oral Health Education (PL-OHE) was adopted as an additional component to the three components of the conventional WHO-BPOC.

Oral Urgent Treatment (OUT) and Atraumatic Restorative Treatment (ART) were provided at schools by qualified dentists once in six months. OUT included; extraction of teeth beyond restorable condition at schools and referral for teeth whose treatment required clinic set up. Whereas ART included; restoring asymptomatic teeth or those in reversible pulpitis state and with cavities accessible by hand instruments. Restorations were done by removing decayed part of a tooth using hand instruments only followed by filling of the cavities with Fuji IX Glass-ionomer cement. Oral health education was provided by trained peers by reading printed messages (box 1) to their classmates once every two weeks.

#### Box 1

**Oral health messages**

1. Restrict the frequency of taking sugary food and drinks in the diet to less than five times per day.
2. Brush your teeth for 2 minutes ensuring all the surfaces are cleaned; twice per day in the morning and evening before retiring to bed.
3. Use fluoride toothpaste to brush, spit the foam but do not rinse it out.
4. Change your toothbrush every 3 months or when bristles flare out.
5. Advise your parent or guardian to buy a tooth paste containing at least 1450 ppm fluoride and to take you for dental checkup at least once a year.

### Control group

The adolescents in the control group were left to continue with their routine daily oral care. Routine daily oral care include all self-oral cleaning and seeking of oral treatment without help from the BPOC project. Participants found with emergence conditions during baseline data collection were referred to nearby dental clinics. At the end of the study, all participants received peer led oral health education based on five key messages (Box 1).

### Study outcome measures

The primary outcome measure of this study was prevalence of dental caries. The study also measured knowledge on dental caries, frequency of consuming sugary drinks and foods per day, use of fluoridated toothpaste per day, and dental visits in the previous year. The primary and secondary outcomes were measured at baseline, at 18 and 24 months follow ups.

## Measurements

### Independent variables

A pre-tested self-administered questionnaire was used to access socio-demographic factors which included age, sex, geographical location, parental education and socioeconomic status. Participant’s age was recorded as an absolute number ranging from 10 to 19, while sex was recoded as 1=male and 2=female. Geographical location was recorded as 1=urban and 2=rural based on Copperbelt province rural-urban delineation. Mother’s and father’s education were recorded according to Zambian education structure [12] as follows: 1=no formal education, 2=up to primary, 3=secondary, 4= tertially (college or university) and thereafter dichotomized into {0=up to primary (including the original scores 1-2) and 1=Secondary or higher (including the original scores 3-4)}. Socio-economic status (SES) was derived from international wealth index (IWI) items [13] as elaborated in a previous publication [14] and recorded as 1= high and 2 =low to middle SES.

### Dependent variables (outcomes)

#### Dental caries

Dental caries on permanent dentition was examined and scored according to categories as described in the CAST manual by Frencken et al., (2015) as follows; 0=sound, 1= sealant, 2= restoration, 3= caries in enamel, 4= caries in dentine without distinct cavitation (discolored dentine visible through enamel), 5= caries in dentine with distinct cavitation, 6= caries in pulp, 7= abscess or fistula, 8= lost due to caries, 9= others [15]. The categories were thereafter grouped into five diagnostic thresholds of CAST as follows; {0=CAST code 0–2 (healthy), 1=CAST code 3–4 (pre-morbidity), 2=CAST code 5 (morbidity), 3=CAST code 6–7 (severe morbidity) and 4= CAST code 8 (mortality). Prevalence of dental caries was assessed by dichotomizing CAST codes into two as “with caries” for CAST codes 3 to7 and “without caries” for CAST codes 0-2, 8, and 9.

### Knowledge and behaviors related to dental caries

Knowledge on dental caries was assessed using ten questions inquiring on causes, symptoms and prevention of oral diseases giving minimum score of 0 and maximum of 10. A cut off point of 7 correct responses was adopted for adequate knowledge; therefore, participants were categorized into those having inadequate knowledge {0 = (including the original scores 0-6)} and having adequate knowledge {1= (including the original scores 7-10)}. Oral health related behaviors assessed were; frequencies of consuming sugary drinks and foods per day in the past 30 days, frequency of use of fluoridated toothpaste per day in the past 30 days, and dental visits in the previous year. Frequency of consuming sugary drinks and foods were scored as {1=I didn’t take, 2=Occasionally per week, 3=Once per day, 4=Twice to four times per day, 5=Five times or more per day} and thereafter dichotomized into {0=less than 5 times per day (including the original scores 1-4)} and 1=5 times or more per day (including the original category 5)}. Frequency of use of fluoridated toothpaste per day was scored as; {1=I didn’t, 2=I did but not every day, 3=I did once a day, 4=I did twice a day or more} and later dichotomized into {0=less than twice per day (including original scores 1-3)} and 1= twice or more per day (including original score 4)}. Visiting a dentist in the previous year was scored as {1=I didn’t attend, 2=I attended once, 3=I attended twice or more} and thereafter dichotomized into {0=I didn’t attend (including original score 1) and 1=I attended once or more (including original score 2-3)}.

### Examination

Dental caries examination was done as prescribed in Caries Assessment Spectrum and Treatment (CAST) manual [16] by four trained and calibrated dentists. A detailed description of examination of dental caries is provided in the article reporting prevalence and factors associated with dental caries among this group [17]. The coefficients of reliability (Cohens kappa) for CAST between and within examiners ranged from 0.80 to 0.90 and 0.80 to 0.85, respectively.

### Sample size and sampling

A sample size of 1,760 participants was estimated by assuming: a 95% confidence level, 85% power, 5% margin of error, 20% expected mean change in dental caries, 0.001 inter-cluster correlation, a cluster size of 80 and a mean DMFT of 1.34 found in a previous study invoving 5 to 17 years old pupils in Zambia [18]. A multistage sampling method was employed to select adolescents in grades 8 and 9 attending 22 public secondary schools in three randomly selected districts of the Copperbelt province in Zambia. The Copperbelt Province was conveniently selected out of the ten provinces of Zambia at the first stage. The choice of Copperbelt Province was based on the intention to fit the study into existing community dentistry training at the university where the principal investigator is based. The second stage involved proportionate stratified random sampling of three districts (one urban and two rural) out of the ten districts of the Copperbelt province. In the third stage, a total of 22 out of 35 public secondary schools were randomly selected guided by the number of schools in each district (21:35, =13 schools for Ndola district; 8:35, = 5 schools for Masaiti district; and 6:35, = 4 schools for Mpongwe district). In the fourth stage, all grade 8 and 9 adolescents enrolled in selected schools were eligible to participate in the study.

### Randomization

The unit of randomization was secondary schools. The allocation ratio of 1:1 resulted into inclusion of 11 schools in the intervention group (908 participants) and 11 in the control group (886 participants) at baseline (Fig 1). Stratified randomization was done in order to prevent imbalance in the rural -urban characteristics of the clusters. Separate randomization was done for urban district (Ndola), and rural districts (Masaiti and Mpongwe). Ndola (urban) clusters were first randomized in a block of 2 by taking the first cluster to intervention and second to control. After completing the Ndola clusters, Masaiti and Mpongwe were randomized in the block size of 2. Sealed envelope [19] was used to create blocked randomization (Seed =72084789304093, block size=2, list length Ndola 13, Actual list length Ndola= 14, List length for Masaiti and Mpongwe = 9, Actual list length for Masaiti and Mpongwe =10). The opaque envelops were opened a day before intervention day. Randomization ended with 7 clusters in intervention group and 6 in control for urban district (Ndola) and 4 clusters in intervention and

5 in control for rural districts (Masaiti and Mpongwe). The Principal Investigator (SA) generated allocation sequence, four dentists enrolled participants and an independent person assigned clusters into interventions.

### Blinding

Single blinding was done whereby the examiners were not aware of group allocation during baseline and follow-up. Blinding of the participants was not feasible because the participants in intervention group received peer delivered oral health education and treatment for dental caries.

#### Statistical methods

Data entry, cleaning and analysis were done using IBM SPSS (version 26.0 IBM SPSS Statistics) and significance level was set to p<0.05. Analysis was based on intention-to-treat approach and generalized estimating equation (GEE) analysis was used to include all available data. The baseline, first and second follow up socio-demographics and clinical characteristics of the participants in the intervention and control groups are presented in frequency tables as percentages and frequencies. The patterns in socio-demographic distribution of outcome variables across the three time points in intervention and control groups are also presented in a frequency table as percentages and frequencies. Chi-squire test was used to ascertain statistically significant differences in the proportions of participants with caries in the intervention and control across baseline, first and second follow up. To take into account of correlated clinical observations within individuals due to repeated measurements, the likelihood of change in caries across study groups was analyzed using Generalized estimating equations (GEE) with binomial model, logit link function and unstructured correlation structure [20]. First GEE analyses were used to analyze the overall intervention effect of the outcome variables and secondly, interactions between intervention/control group and time were explored to evaluate the intervention effects at follow up and whether the outcome variables showed different trajectories across time. Sub group analyses were performed for the outcome variables which showed interaction effects with time. Results of GEE are presented as adjusted odds ratios and their 95% confidence intervals.

## Results

### Participants flow diagram

The participants flow diagram of this study is illustrated in Figure 1. A total of 35 schools from Ndola, Masaiti and Mpongwe districts of the Copperbelt province were eligible to participate in this trial during the enrollment period (February-May 2021). Out of the 35 schools, one school declined to participate resulting into 34 schools being available for random selection. Using computer generated random numbers 22 out of 34 schools were randomly selected. All adolescents in grades 8 and 9 meeting inclusion criteria and were available during enrollment period were invited to participate.

A total of 115 students were either excluded due to being under orthodontic treatment (2) or declined to participate (113). The twenty-two schools were allocated randomly into intervention and control in a ratio of 1:1 resulting in 908 and 886 participants in intervention and control groups, respectively. Intervention group received modified BPOC from June to December 2021 while control group did not receive any intervention and therefore continued with their routine oral care. There was no complete loss to follow up in any of the clusters at the two follow ups. A total of 64 participants in the intervention and 40 in the control group were lost to follow up after 18 months while 117 participants in the intervention and 80 in the control group were lost to follow-up after 24 months; all of them due to being transferred from one school to another.

A total of 1,794 participants (908-intervention, 886-control) and 1,597 participants (791-intervention, 806-control) were analyzed at baseline and after 24 months, respectively. The trial ended in March 2023.

### Baseline characteristics

Baseline socio-demographics, oral health related knowledge and behaviors, and clinical indicators of the participants according to trial arm allocation are as shown in table 1. Despite randomization chi-square test and variance analysis showed significant baseline differences between the intervention and control arms on the following socio-demographic characteristics; sex (p=0.003), mothers’ education (p=0.023), fathers’ education (p= 0.021), and socioeconomic status (p <0.001). The intervention group had higher proportion of females (57.4% vs 50.5%), rural participants (56.8% vs 33.2%), participants whose mothers (38.4% vs 33.3%) and fathers (43.4% vs 38.0%) had attained secondary school or higher education and low to middle SES (52% vs 35.3%) compared to controls. The results also show baseline difference according to dental visit in previous one year (p=0.004), and CAST clinical severity categories (p<0.001). The intervention group had significantly higher percentages of those who did not attend for dental visits (76.5% vs 70.5%), and those with dental caries (51.3 % vs 41.3%) than control group. All the effect sizes of the variances were small according to Ferguson, (2009) [21].

**Table 1:**
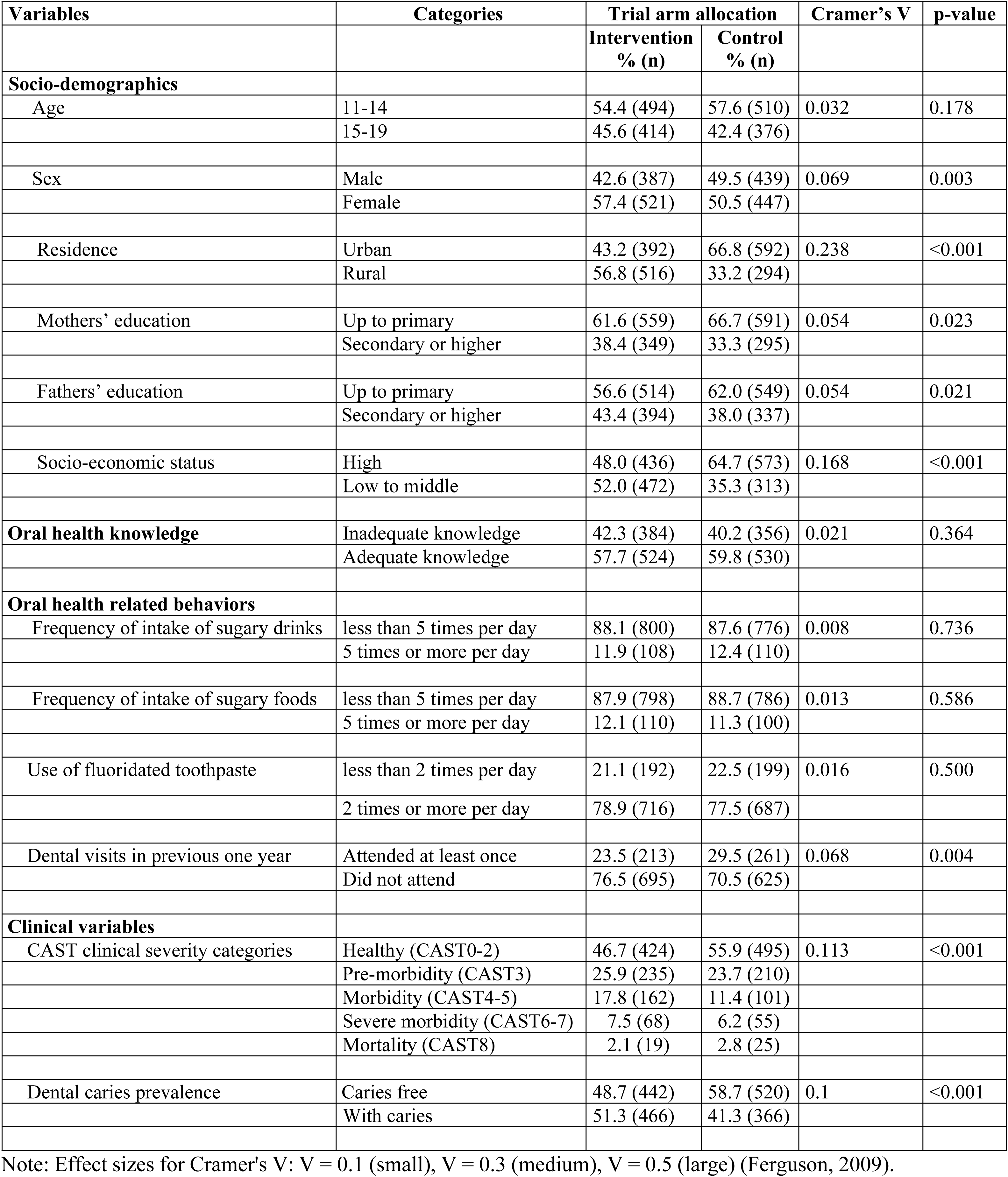
Socio-demographics, oral health knowledge, oral health behaviors and clinical characteristics by intervention arms at baseline (n=1794)

Table 2 shows socio-demographics and clinical variables at baseline by response status at 18- and 24months follow up. The baseline socio-demographics (age, sex, residence, mothers’ education, fathers’ education), oral health related knowledge and behaviors and clinical variables (caries) between those lost to follow up and those who remained in the study did not differ significantly (all p-values >0.05).

**Table 2:** Socio-demographics and clinical variables at baseline by response status.

| Variables | Categories | Follow ups |  |  |  |  |  |
| --- | --- | --- | --- | --- | --- | --- | --- |
|  |  | 18 months |  |  | 24 months |  |  |
|  |  | Lost<br>% (n) | Followed<br>% (n) | p<br>value | Lost<br>% (n) | Followed<br>% (n) | p<br>value |
| **Socio-demographics** |  |  |  |  |  |  |  |
| Age | 11-14 | 67.3 (70) | 55.3 (934) | 0.016 | 59.4 (117) | 55.5 (887) | 0.305 |
|  | 15-19 | 32.7 (34) | 44.7 (756) |  | 40.6 (80) | 44.5 (710) |  |
| Sex | Male | 48.1 (50) | 45.9 (776) | 0.668 | 44.7 (88) | 46.2 (738) | 0.682 |
|  | Female | 51.9 (54) | 54.1 (914) |  | 55.3 (109) | 53.8 (859) |  |
| Residence | Urban | 58.7 (61) | 54.6 (923) | 0.422 | 53.8 (106) | 55.0 (878) | 0.755 |
|  | Rural | 41.3 (91) | 45.4 (767) |  | 46.2 (91) | 45.0 (719) |  |
| Mothers' education | Up to primary | 65.4 (68) | 64.0(1082) | 0.779 | 62.9 (124) | 64.2(1026) | 0.719 |
|  | Secondary or higher | 34.6 (36) | 36.0 (608) |  | 37.1 (73) | 35.8 (571) |  |
| Fathers' education | Up to primary | 66.3 (69) | 58.8 (994) | 0.129 | 62.9 (124) | 58.8 (939) | 0.264 |
|  | Secondary or higher | 33.7 (35) | 41.2 (696) |  | 37.1 (73) | 41.2 (658) |  |
| Socio-economic status | High | 65.4 (68) | 55.7 (941) | 0.053 | 60.4 (119) | 55.7 (890) | 0.212 |
|  | Low to middle | 34.6 (36) | 44.3 (749) |  | 39.6 (78) | 44.3 (707) |  |
| Oral health knowledge | Inadequate | 38.5 (40) | 41.4 (700) | 0.552 | 41.6 (82) | 41.2 (658) | 0.910 |
|  | Adequate knowledge | 61.5 (64) | 58.6 (990) |  | 58.4 (115) | 58.8 (939) |  |
| **Oral health related behaviors** |  |  |  |  |  |  |  |
| Frequency of intake of sugary drinks | less than 5 times per day | 88.5 (92) | 87.8 (1484) | 0.844 | 88.3 (174) | 87.8 (1402) | 0.828 |
|  | 5 times or more per day | 11.5 (12) | 12.2 (206) |  | 11.7 (23) | 12.2 (195) |  |
| Frequency of intake of sugary foods | less than 5 times per day | 82.7 (86) | 88.6 (1498) | 0.067 | 87.3 (172) | 88.4 (1412) | 0.649 |
|  | 5 times or more per day | 17.3 (18) | 11.4 (192) |  | 12.7 (25) | 11.6 (185) |  |
| Use of fluoridated toothpaste | less than 2 times per day | 16.3 (17) | 22.1 (374) | 0.166 | 19.8 (39) | 22.0 (352) | 0.472 |
|  | 2 times or more per day | 83.7 (87) | 77.9 (1316) |  | 80.2 (158) | 78.0 (1245) |  |
| Dental visits in previous one year | Attended at least once | 26.0 (27) | 26.4 (447) |  | 26.9 (53) | 26.4 (421) | 0.871 |
|  | Did not attend | 74.0 (77) | 73.6 (1243) | 0.871 | 73.1 (144) | 73.6 (1176) |  |
| **Clinical variables** |  |  |  |  |  |  |  |
| CAST clinical severity categories | Healthy (CAST0-2) | 51.9 (54) | 51.2 (865) | 0.853 | 52.3 (103) | 51.1 (816) | 0.750 |
|  | Pre-morbidity (CAST3) | 25.0 (26) | 24.8 (419) |  | 27.4 (54) | 24.5 (391) |  |
|  | Morbidity (CAST4-5) | 11.5 (12) | 14.9 (251) |  | 12.7 (25) | 14.9 (238) |  |
|  | Severe morbidity (CAST6-7) | 8.7 (9) | 6.7 (114) |  | 5.6 (11) | 7.0 (112) |  |
|  | Mortality (CAST8) | 2.9 (3) | 2.4 (41) |  | 2.0 (4) | 2.5 (40) |  |
| Dental caries prevalence | Caries free | 54.8 (57) | 53.6 (905) | 0.803 | 54.3 (107) | 53.5 (855) | 0.837 |
|  | With caries | 45.2 (47) | 46.4 (785) |  | 45.7 (90) | 46.5 (742) |  |

### Trends in the distributions of participants’ knowledge, oral health related behaviors and dental caries prevalence by trial arms and across time

Table 3 shows the trends in the distributions of participants according to knowledge, oral health related behaviors and dental caries by intervention group and across time. The results show no baseline differences (p=0.364) in knowledge between intervention and control groups. However, follow up results show statistically significant difference between groups after 18 months (p=0.014) and 24 months (p=0.002). The percentage change in the proportion of participants with adequate knowledge from baseline to 18-and-24 months follow up was significant in the intervention {9.6% (5.1%, 14.1%), p<0.001} at 18 months and 11.5% (5.1%, 14.1%), p<0.001) at 24 months while the change was not significant in the control group {1.8% (-2.8%, 6.4%), p=0.443} at 18 months and {2.1% (-2.6%, 6.7%), p=0.377} at 24 months. The use of fluoridated toothpaste in the intervention group increased by 4.5% (0.8%, 8.1%), p=0.016 and 4.4 % (0.6%, 8.1%), p=0.021 at 18 and 24 months respectively. On the other hand, the results in the controls showed no significant change at 18 {-0.4% (-3.5%, 4.4%), p=0.843} and 24 months {-0.2% (-3.8%, 4.2%), p=0.922}, respectively. Frequency of dental visits was lower in intervention than control group (23.5% vs 29.5%, p<0.001) at baseline but changed to no difference at 18 months (p=0.733) and 24 months (p=0.688). Dental visit in the intervention group increased by 5.6% {5.6% (1.5%, 9.7%) p=0.008} and 5.4 % {5.4% (1.2%, 9.6%) p=0.011} at 18 and 24 months respectively while no significant change was observed in the control group at 18 months {0.4% (-3.9%, 4.7%) p=0.856} and at 24 months {0.3% (-4.1%, 4.7%) p=0.893}. The prevalence of dental caries was significantly higher in the intervention group than in the control group at baseline, 18 months and 24 months (p<0.001). The prevalence of dental caries from baseline to 18 months’ follow-up and from baseline to 24 months’ follow-up did not change significantly either in the intervention-nor in the control groups. The percentage change, 95% confidence interval and p values for the intervention at 18 months and 24 months were {-2.5% (-2.2%, 7.2%) p=0.296}and {-2.6% (-2.2%, 7.3%) p=0.285} respectively. The corresponding change estimates in the control group were { -0.9 (-3.7, 5.5), p=0.703} at 18 months and { -2.3 (-2.3, 6.9), p=0.335} at 24 months.

**Table 3.** Distributions of participants according to knowledge, oral health related behaviors and dental caries by trail arm across time.

| Outcome variable | Percentage and number across time |  |  | Percentage change from baseline (95%CI) and p value |  |
| --- | --- | --- | --- | --- | --- |
|  | Baseline % (n) | 18 months % (n) | 24 months % (n) | at 18 months % Δ (95%CI), p value | at 24 months % Δ (95%CI), p value |
| **Knowledge (adequate)** |  |  |  |  |  |
| intervention | 57.7 (524) | 67.3 (568) * | 69.2 (547) * | 9.6 (5.1, 14.1), p<0.001 | 11.5 (6.9, 15.9), p<0.001 |
| control | 59.8 (530) | 61.6 (521) | 61.9 (499) | 1.8 (-2.8, 6.4), p=0.443 | 2.1 (-2.6, 6.7), p=0.377 |
| **Oral health behaviors** |  |  |  |  |  |
| Sugary drinks per day (less than 5 times) |  |  |  |  |  |
| intervention | 88.1 (800) | 87.6 (739) | 87.2 (689) | -0.5 (-2.5, 3.6), p=0.749 | -0.9 (-2.2, 4.1), p=0.573 |
| control | 87.6 (776) | 86.9 (735) | 86.6 (698) | -0.7 (-2.4, 3.9), p=0.662 | 1.0 (-2.2, 4.2), p=0.539 |
| Sugary foods per day (less than 5 times) |  |  |  |  |  |
| intervention | 87.9 (798) | 87.7 (740) | 87.3 (690) | -0.2 (-2.9, 3.3), p=0.898 | -0.6 (-2.6, 3.8), p=0.708 |
| control | 88.7 (786) | 88.8 (751) | 88.3 (712) | 0.1 (-2.9, 3.1), p=0.947 | -0.4 (-2.6, 3.5), p=0.797 |
| Fluoridated toothpaste per day (2 times/more) |  |  |  |  |  |
| intervention | 78.9 (716) | 83.4 (704) * | 83.3 (658) * | 4.5 (0.8, 8.1), p=0.016 | 4.4 (0.6, 8.1), p=0.021 |
| control | 77.5 (687) | 77.1(652) | 77.3 (623) | -0.4 (-3.5, 4.4), p=0.843 | -0.2 (-3.8, 4.2), p=0.922 |
| Dental visits in previous one year (attended) |  |  |  |  |  |
| intervention | 23.5 (213) * | 29.1 (246) | 28.9 (228) | 5.6 (1.5, 9.7), p=0.008 | 5.4 (1.2, 9.6), p=0.011 |
| control | 29.5 (261) | 29.9 (253) | 29.8 (240) | 0.4 (-3.9, 4.7), p=0.856 | 0.3 (-4.1, 4.7), p=0.893 |
| **Dental caries (with caries)** |  |  |  |  |  |
| intervention | 51.3 (466) ** | 48.8 (412) * | 48.7 (385) * | -2.5 (-2.2, 7.2), p=0.296 | -2.6 (-2.2, 7.3), p=0.285 |
| control | 41.3 (366) | 40.4 (342) | 39.0 (314) | -0.9 (-3.7, 5.5), p=0.703 | -2.3 (-2.3, 6.9), p=0.335 |

Percentage and numbers given are for only one category of the outcome variable as indicated in bracket in front of the outcome variable, \*=p value <0.05, \*\*=p<0.001, Δ=percentage

Figure 2 illustrates percentage distribution of dental caries according to CAST clinical categories by trial arms at baseline (T_1_), first (T_2_) and second (T_3_) follow up. Generally, the percentages of participants decreased as the severity of CAST clinical categories increased for both intervention and control arms. CAST 0-2 and CAST 6-7 increased while CAST 5 decreased in both trial arms. There was an increase in the mortality stage (CAST 8) across time in both arms however, the increase was higher in the control than intervention arm.

### Effects of the BPOC intervention on outcomes (Generalized estimating equations) at 18 months

Generalized estimating equation models were used to assess the effect of the BPOC intervention at 18 months follow up regarding the outcome variables (knowledge, oral health behaviors and dental caries prevalence) as shown in table 4. At 18 months, a significant interaction effect between time and group on adequate knowledge across time occurred after adjustment for the main effects of group and time (BPOC group x time interaction) {OR (95%CI) = 1.5 (1.2,1.8), p<0.001}. The model for use of fluoridated toothpaste twice or more per day also showed significant interaction between BPOC group and time {OR (95%CI) = 1.6 (1.3, 2.1), p<0.001)}. Dental caries interaction model at 18 months was also significant {OR (95%CI) = 1.3 (1.1, 1.6), p=0.003)}. There were no significant interactions between BPOC intervention and time at 18 months for frequency of intake of sugary drinks of less than 5 times per day model (p=0.506), frequency of intake of sugary foods of less than 5 times per day foods model (p=0.505) and at least once per year dental visit model (p=0.422).

**Table 4.** Generalized estimating equation analyses of the outcome variables (Knowledge, behaviors and dental caries) for the study sample from baseline to 18 months.

| Outcome variable | Parameters | Estimate (SE) | Sig. | OR (95%CI) |
| --- | --- | --- | --- | --- |
| **Adequate Knowledge** |  |  |  |  |
| Main effects | Time | 0.274 (0.035) | < 0.001 | 1.3 (1.2, 1.4) |
|  | BPOC group | 0.134 (0.094) | 0.154 | 1.1 (1.0, 1.4) |
| Interaction effects | BPOC group x time | 0.392 (0.102) | < 0.001 | 1.5 (1.2, 1.8) |
| **Behaviors** |  |  |  |  |
| Sugary drinks per day (less than 5 times) |  |  |  |  |
| Main effects | Time | -0.062 (0.049) | 0.207 | 0.9 (0.8, 1.0) |
|  | BPOC group | -0.037 (0.139) | 0.790 | 1.0 (0.7, 1.3) |
| Interaction effects | BPOC group x time | -0.098 (0.148) | 0.506 | 0.9 (0.7, 1.2) |
| Sugary foods per day (less than 5 time) |  |  |  |  |
| Main effects | Time | -0.007 (0.046) | 0.873 | 1.0 (0.8, 1.1) |
|  | BPOC group | -0.092 (0.142) | 0.515 | 0.9 (0.7, 1.2) |
| Interaction effects | BPOC group x time | -0.100 (0.149) | 0.505 | 0.9 (0.7, 1.2) |
| Fluoridated toothpaste per day (2 times/more) |  |  |  |  |
| Main effects | Time | 0.134 (0.041) | 0.001 | 1.1 (1.1, 1.3) |
|  | BPOC group | 0.329 (0.114) | 0.004 | 1.4 (1.1, 1.7) |
| Interaction effects | BPOC group x time | 0.485 (0.125) | < 0.001 | 1.6 (1.3, 2.1) |
| Dental visits in previous one year (attended) |  |  |  |  |
| Main effects | Time | 0.157 (0.035) | < 0.001 | 1.2 (1.1, 1.3) |
|  | BPOC group | -0.074 (0.102) | 0.470 | 0.9 (0.8, 1.1) |
| Interaction effects | BPOC group x time | 0.086 (0.108) | 0.422 | 1.1 (0.9, 1.4) |
| **Dental caries (with caries)** |  |  |  |  |
| Main effects | Time | -0.070 (0.025) | < 0.001 | 0.9 (0.8, 0.9) |
|  | BPOC group | 0.359 (0.094) | < 0.001 | 1.4 (1.2, 1.7) |
| Interaction effects | BPOC group x time | 0.289 (0.098) | 0.003 | 1.3 (1.1, 1.6) |

Abbreviations: SE = Standard error, OR = Odds ratio, CI= confidence intervals, Sig.= significancy, BPOC –
Basic package of oral care. Note: results provided are based on intervention group and at 18 months with reference to control group and baseline data

### Effects of the BPOC intervention on outcomes (Generalized estimating equations) at 24 months

Table 5 shows the results from GEE models regarding the effect of the BPOC intervention at 24 months follow -up. The intervention group (BPOC group x time interaction) for adequate knowledge model was significant at 24 months {OR (95%CI) = 1.6 (1.3, 2.0)}, indicating group different trajectories for knowledge across time from baseline to 24 months follow-up. The model for use of fluoridated toothpaste twice or more per day also showed significant interaction between BPOC group and time {OR (95%CI) = 1.6 (1.3, 2.1), p<0.001)} at 24 months. Dental caries interaction model at 24 months was significant {OR (95%CI) = 1.3 (1.1, 1.6), p=0.004)}.

**Table 5.** Generalized estimating equation analyses of the outcome variables (Knowledge, behaviors and dental caries) for the study sample from baseline to 24 months.

| Outcome variable | Parameters | Estimate (SE) | Sig. | OR (95%CI) |
| --- | --- | --- | --- | --- |
| **Adequate Knowledge** |  |  |  |  |
| Main effects | Time | 0.292 (0.036) | < 0.001 | 1.3 (1.2, 1.4) |
|  | BPOC group | 0.131 (0.094) | 0.162 | 1.1 (0.9, 1.4) |
| Interaction effects | BPOC group x time | 0.476 (0.103) | < 0.001 | 1.6 (1.3, 2.0) |
| **Behaviors** |  |  |  |  |
| Sugary drinks per day (less than 5 times) |  |  |  |  |
| Main effects | Time | -0.082 (0.048) | 0.089 | 0.9 (0.8, 1.0) |
|  | BPOC group | -0.042 (0.139) | 0.764 | 0.9 (0.7, 1.3) |
| Interaction effects | BPOC group x time | -0.124 (0.146) | 0.394 | 0.9 (0.7, 1.2) |
| Sugary foods per day (less than 5 time) |  |  |  |  |
| Main effects | Time | -0.057 (0.043) | 0.180 | 0.9 (0.8, 1.0) |
|  | BPOC group | -0.090 (0.140) | 0.520 | 0.9 (0.7, 1.2) |
| Interaction effects | BPOC group x time | -0.147 (0.146) | 0.317 | 0.8 (0.6, 1.2) |
| Fluoridated toothpaste per day (2 times/more) |  |  |  |  |
| Main effects | Time | 0.147 (0.040) | < 0.001 | 1.2 (1.1, 1.3) |
|  | BPOC group | 0.287 (0.113) | 0.011 | 1.3 (1.1, 1.7) |
| Interaction effects | BPOC group x time | 0.490 (0.124) | < 0.001 | 1.6 (1.3, 2.1) |
| Dental visits in previous one year (attended) |  |  |  |  |
| Main effects | Time | 0.149 (0.034) | < 0.001 | 1.2 (1.1, 1.3) |
|  | BPOC group | -0.092 (0.103) | 0.371 | 0.9 (0.7, 1.1) |
| Interaction effects | BPOC group x time | 0.057(0.107) | 0.596 | 1.1 (0.9, 1.3) |
| **Dental caries (with caries)** |  |  |  |  |
| Main effects | Time | -0.107 (0.028) | < 0.001 | 0.9 (0.8, 0.9) |
|  | BPOC group | 0.384 (0.093) | < 0.001 | 1.5 (1.2, 1.8) |
| Interaction effects | BPOC group x time | 0.276 (0.097) | 0.004 | 1.3 (1.1, 1.6) |

Abbreviations: SE = Standard error, OR = Odds ratio, CI= confidence intervals, Sig.= significancy, BPOC – Basic package of oral care. Note: results provided are based on intervention group and at 24 months with reference to control group and baseline data

### Subgroup analyses

Subgroup analyses of the outcome variables with significant interactions between BPOC group and time at 18 and 24 months are as shown in table 6. The findings show that the likelihoods of having adequate knowledge at 18- and 24 months follow ups compared to baseline were stronger in the intervention than in the control group. The odds ratio, 95% confidence intervals and p values for adequate knowledge at 18 months were {OR (95%CI) = 1.5 (1.4, 1.7), p<0.001)} for intervention and {OR (95%CI) = 1.1 (1.0, 1.2), p=0.020)} control group compared to baseline values. The odds ratios, 95% confidence intervals and p values for intervention and control group at 24 months compared to baseline values were {OR (95%CI) = 1.6 (1.5, 1.9), p<0.001)} and {OR (95%CI) = 1.1 (1.0, 1.2), p=0.020)} respectively. The likelihood of using fluoridated toothpaste two times or more per day at 18 and 24 months of follow up compared to baseline was stronger in the intervention than in the control group. The use of fluoridated toothpaste increased by about 1.4 times at both 18 months {OR (95%CI) = 1.4 (1.2, 1.6), p<0.001)} and 24 months {OR (95%CI) = 1.4 (1.2, 1.5), p<0.001)} in the intervention group but did not change significantly for control group. The likelihood of having dental caries at 18 and 24 months follow up compared to baseline decreased more in the intervention than in the control group. The prevalence of dental caries in the intervention group decreased by 20% at both 18 months {OR (95%CI) = 0.8 (0.8, 0.9), p=0.013)} and 24 months {OR (95%CI) = 0.8 (0.8, 0.9), p=0.021)} compared to 10% decrease in the control group at 18 months {OR (95%CI) = 0.9 (0.8, 0.9), p=0.002)} and 24 months {OR (95%CI) = 0.9 (0.8, 0.9), p<0.001)}.

**Table 6.**
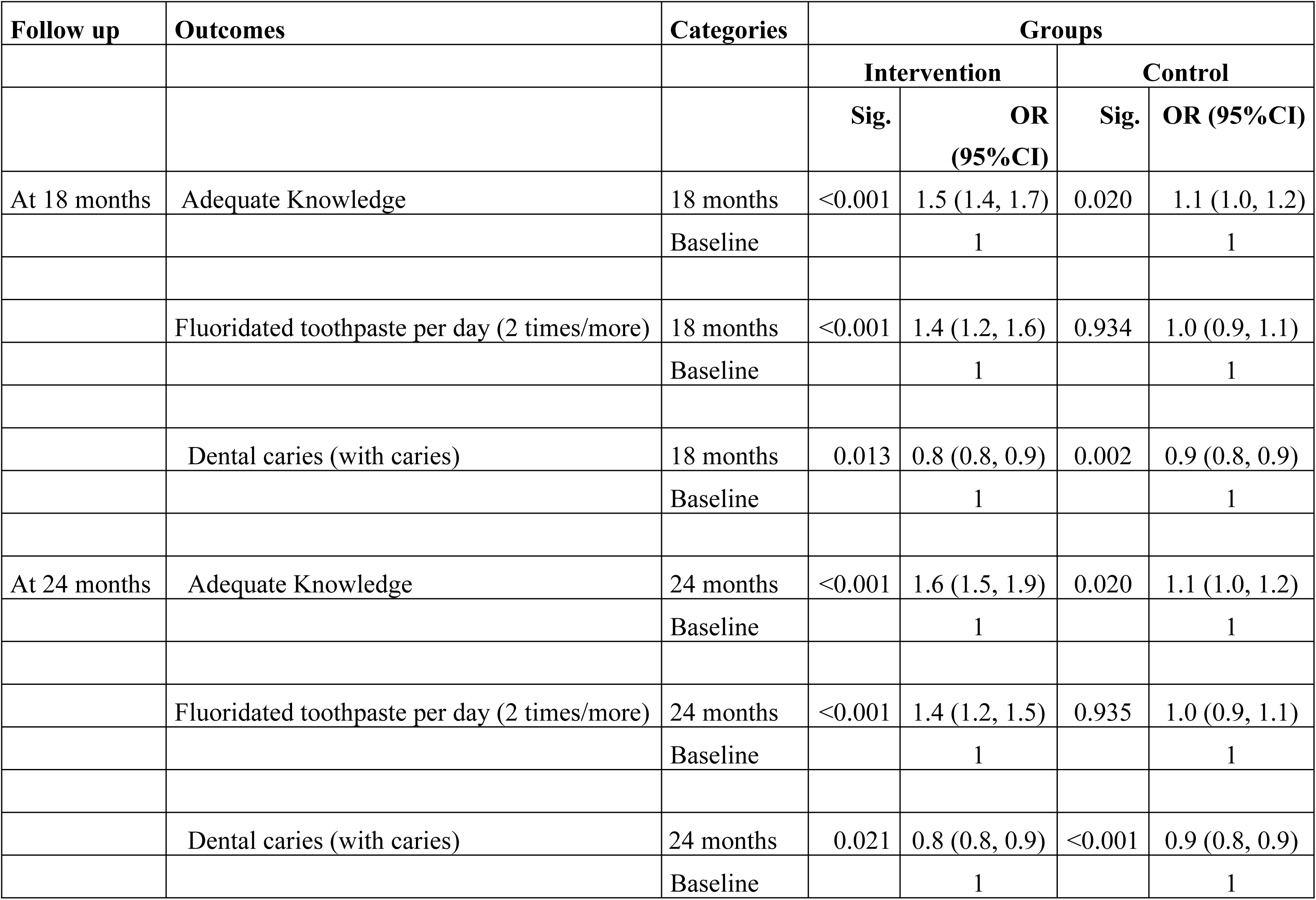
BPOC outcome variables regressed on time (18 – and 24 months follow up versus baseline) separately in the intervention and control groups.

## Discussion

This study investigated the impact of a modified basic package of oral care intervention on prevalence of dental caries as well as knowledge and oral health behaviors related to dental caries among adolescents in Copperbelt Province, Zambia.

The major findings of the current study indicate that at both six and twelve months after cessation of BPOC intervention, knowledge on dental caries and frequency of using fluoridated toothpastes among participants in BPOC group improved over time. The frequency of consumption of sugary drinks and foods did not significantly change over time for either the intervention or control group. Over time, the prevalence of dental caries decreased in both groups; however, the intervention group had a twofold decrease in dental caries compared to the control group.

Improvement in knowledge among the participants in the BPOC group was likely attributed to repetitive oral health messages delivered by peers. Comparable improvements in knowledge have been reported in previous studies which used peer-led oral health education [22–24].

Surprisingly, the observed positive change in knowledge in the BPOC group in this study did not lead to favorable behavior change over time The lack of favorable change in the consumption of sugary foods and drinks despite improvement of knowledge could be attributed to immaturity in the brain pathways involved in decision making, making adolescents sensitive to emotional and social influences and therefore difficulty to control their lifestyle choices [25]. Adolescents are also dependents, thus their eating behaviours are not only influenced by their knowldge but also by what foods are available and accessible to them, which ultimately influences what they consume [26]. This finding hightlights the impact of other determinants of behaviours such as the environment, cultural and social norms, social support from friends (peers) and parents that influence behaviour change, which were not evaluated or controlled in this cluster randomized trial (27). In contrast to the current findings, other authors [23,24,28] have reported positive change in both knowledge and behavior following peer oral health education.

Participants in the BPOC group used fluoridated toothpaste more frequently than controls in this study. This could be attributed not only to increased knowledge about dental caries which was given through peer-led OHE but also by a motivation to continue using oral cleaning products after receiving free toothpaste during the six-month intervention period. The importance of knowledge, particularly the benefits of fluoride use in caries prevention, constitutes one of the major factors influencing adolescents’ use of fluoridated toothpaste [29].

The more significant improvement in the intervention group than the control group, regarding usage of fluoridated toothpaste is comparable with the findings of a previous trial in which free toothpaste and toothbrushes were distributed along with messages and instructions on daily brushing [30].

The treatment components of the BPOC intervention which included extraction of teeth and atraumatic restorative treatment could explain the two-fold decrease in the prevalence of dental caries among participants in BPOC compared to control group. Frequent use of fluoridated toothpaste in the BPOC group compared to controls may also have contributed to higher improvement in caries status in the BPOC group. Comparison with studies on the effectiveness of BPOC intervention in caries prevention among adolescents was not possible due to scarcity of similar studies. However, the findings are consistent with those of a study that investigated the effectiveness of oral urgent treatment, one of the four components of BPOC, in lowering the dental caries burden in Filipino children [31].

The findings of this study need to be interpreted within the strengths and limitations of a cluster randomized trial. The study involved a large sample size from a total of twenty-two clusters which exceed the recommended minimum number of clusters for a parallel randomized cluster trials of four per arm in order to obtain a p-value less than 0.05 under a randomization-based test [32]. The findings can probably be generalized to adolescents attending public secondary schools in Copperbelt Province, Zambia, due to random selection and a large sample size. The design employed in this study allowed prevention of contamination of the intervention which could lead to false rejection of an effective intervention due to reduction of point estimates of its effectiveness [33]. Data collection assistants were blinded in order to avoid bias in their assessment of dental caries status which could ultimately affect assessment of the effectiveness of an intervention [34]. However, it was not possible to blind the participants due to the nature of the BPOC intervention which involved peer oral education, provision of fluoridated toothpastes, extraction and restoration of teeth. It is also important to note that despite randomization there was baseline differences in dental caries and dental visits as well as in some socio-demographics between intervention and control group which could affect treatment effects of the BPOC intervention. Use of a repeated measure (GEE) without the treatment variable but interaction between the treatment variable and time in the models has been used to overcome this shortcoming (35). The short follow-up period and age group did not allow for a proper evaluation of the preventive effect of affordable fluoride and atraumatic restorative treatment.

## Conclusion

Basic package of oral care was effective in reducing prevalence of dental caries, improving knowledge on dental caries, and the frequency of using fluoridated toothpaste among Zambian adolescents. The intervention did not have any effect on adolescent’s frequency of consuming sugary drinks and foods. Further studies need to be conducted in order to address other factors affecting consumption of sugary diet behaviors such as environmental, social and cultural factors.

## Supporting information

S1-CONSORT 2010 checklist of information to include when reporting a randomised trial S2- Schedule of study activities.

## Data Availability

Data cannot be shared publicly but can be accessed from the principal investigator upon request for researchers who meet the criteria for access to confidential data.

## Acknowledgements

We appreciate support from District Education Board Secretaries (DEBS), Head teachers & study participants and Prof Stein Atle Lie (UIB-Norway) for guidance during data analysis.

